# The “Walking for Harm Reduction Through Street Engagement” Social Media Knowledge Translation Strategy

**DOI:** 10.1101/2025.08.01.25332613

**Authors:** Nana Koomson, Muna Aden, Anita C. Benoit

**Author notes:** **Corresponding Author:** Anita C. Benoit, Highland Hall, Room HL230, University of Toronto Scarborough 1265 Military Trial, Scarborough, Ontario, M1C 1A4.

## Abstract

Due to its ability to cost-effectively reach underrepresented groups, social media is a valuable tool for facilitating knowledge translation. Consequently, a Social Media Knowledge Translation Strategy was developed for the Walking for Harm Reduction Through Street Engagement (WHiSE 2.0) study; a research project that seeks to identify the harm reduction approaches desired and used by Indigenous people in Thunder Bay, Sault Ste. Marie, and Sudbury. The strategy aided in the referral of harm reduction services to Indigenous people in the WHiSE 2.0 study sites, build relationships between Indigenous communities and harm reduction organizations, and increase knowledge translation and exchange (KTE) efforts among a target audience. The Social Media Knowledge Translation Strategy was developed following Elliot et al.’s [14] social media knowledge translation stages of planning, doing, and evaluating, and quantitative data was collected from an Instagram and Twitter account between May 2023 and August 2023. The results showed a disproportionate ratio of new followers to reach, content interactions, and profile visits. Additionally, there was higher reach and impressions compared to engagement, and the aspiration of amassing 150 followers on Instagram, Facebook, and Twitter which was not realized. However, as any number of impressions, profile visits, content interactions, reach, engagement, and new followers beyond 0, was viewed as successfully disseminating knowledge to at least one organization or individual, the strategy successfully engaged the intended target audience. In future iterations of the strategy, metrics beyond social media insights must be collected to fully evaluate the success of the strategy.

## INTRODUCTION

### Knowledge Translation

Also referred to as knowledge translation and exchange (KTE), knowledge dissemination, or knowledge mobilization, knowledge translation encompasses the synthesis, dissemination, exchange, and ethical application of research or knowledge [1–3]. The process involves dynamic and iterative interactions between research producers and users, aimed at bridging the gap between knowledge and practice [4–6].

Several knowledge translation models exist, but push, pull, exchange, and integrated strategies are among the most widely recognized [7–9]. Push strategies involve transferring information from researchers, intermediary stakeholders or rights-holders, or knowledge brokers to research users who may not be aware of specific knowledge products or outcomes [7–10]. In contrast, pull mechanisms are driven by research users who actively seek out information and evidence to inform their decision-making processes [7, 9] Exchange models emphasize collaboration and partnerships between research producers and users, promoting a shared understanding of relevant research questions and methodologies [4, 9–10]. Lastly, integrated knowledge translation strategies involve engaging research users—such as policymakers, clinicians, service users, community members, and organizations—throughout the entire research process to develop evidence-based solutions [1, 11–12]. Integrated knowledge translation strategies allow research users to invoke changes during and after the completion of research projects [11].

### Using Social Media for Knowledge Translation

Social media, a user-centered internet-based tools and platforms that enable the creation and sharing of content, is an effective knowledge translation tool by facilitating engagement and interaction among a diverse range of research producers and users across various sectors of society [13–16]. Knowledge translation through social media can occur via various platforms, including blogs, wikis, podcasts, microblogging services like Twitter, social networking platforms such as Facebook and Instagram, and video-sharing sites like YouTube. While leveraging the reach of mass media, it is driven by the social networking capabilities of individuals and organizations [17]. Additionally, it provides a cost-effective means to connect with populations that are often neglected, ignored, or under-represented in health-related research [18].

In the healthcare field, social media can advance knowledge translation by distributing health information, delivering health interventions, fostering networking among health professionals, collecting surveillance data, facilitating health promotion, and building patient communities [15, 19–20]. Due to its capacity for wide distribution, promotion of audience-specific messages, and support for community building and advocacy, social media is increasingly integrated into various public health interventions [19]. Unlike academic journals, information shared through social media is more accessible, efficient for research users, engaging, and easier to understand. It also allows for faster dissemination and improved two-way communication between research producers and users [15].

My paper examines how a Social Media Knowledge Translation Strategy was created to disseminate information on harm reduction, knowledge translation, and community-based research to Indigenous communities as part of the Walking for Harm Reduction through Street Engagement project. Social media was selected for knowledge translation due to its potential to bridge gaps that often exist between health research and practice in Indigenous communities. Despite frequent neglect of such translation activities, some authors suggest that social media can effectively engage these communities [21–22].

### Walking for Harm Reduction Through Street Engagement (WHiSE) 2.0

WHiSE 2.0 is a three-year research study that seeks to identify the harm reduction approaches desired and used by Indigenous people in Thunder Bay, Sault Ste. Marie, and Sudbury, Ontario. Collecting data on harm reduction will allow the research team to assess the success of harm reduction strategies by Elevate NWO and the Ontario Aboriginal HIV/AIDS Strategy (Oahas) staff to optimize their services to better meet the needs of their Indigenous clients.

The purpose of this 4-month social media development plan is to address the three KTE objectives of WHiSE 2.0: 1) aid in the referral and delivery of harm reduction services to Indigenous people in Thunder Bay, Sault Ste. Marie, and Sudbury; 2) build relationships between Indigenous communities and harm reduction organizations; and 3) increase KTE knowledge and strategies among different target audience.

## SOCIAL MEDIA KNOWLEDGE TRANSLATION STRATEGY

### Social Media Knowledge Translation Framework

Elliot et al. described planning, doing, and evaluating as factors to consider when implementing a successful Social Media Knowledge Translation Strategy. Planning refers to the advanced planning of specific elements required to undertake a Social Media Knowledge Translation Strategy. This includes a prior understanding of the best platforms to use, the scope of the messaging, strategies to gather knowledge from sources, and strategies to engage with knowledge users. Doing refers to the activities undertaken to complete the Social Media Knowledge Translation Strategy. Such activities include building an audience and engaging with them on identified social media platforms and creating compelling digital content. Lastly, evaluating indicates the strategies used to understand how different facets of the Social Media Knowledge Translation Strategy performed [14].

The Social Media Knowledge Translation Strategy employed for the WHiSE 2.0 study aligns closely with planning, doing, and evaluating. Before embarking on the framework, the goals of the Social Media Knowledge Translation Strategy were established and connected to relevant objectives of the WHiSE 2.0 study. As evident in Table 1, this was to ensure that the process of planning, doing, and evaluating were structured according to specific goals and objectives. The identified social media goals include developing a target audience, increasing awareness of harm reduction which includes substance use disorder support services in Thunder Bay, Sault Ste Marie, and Sudbury, improving community understanding of harm reduction, increasing engagement with harm reduction services, creating conversations, and increasing understanding of knowledge translation and engagement theories and strategies.

**TABLE 1.**
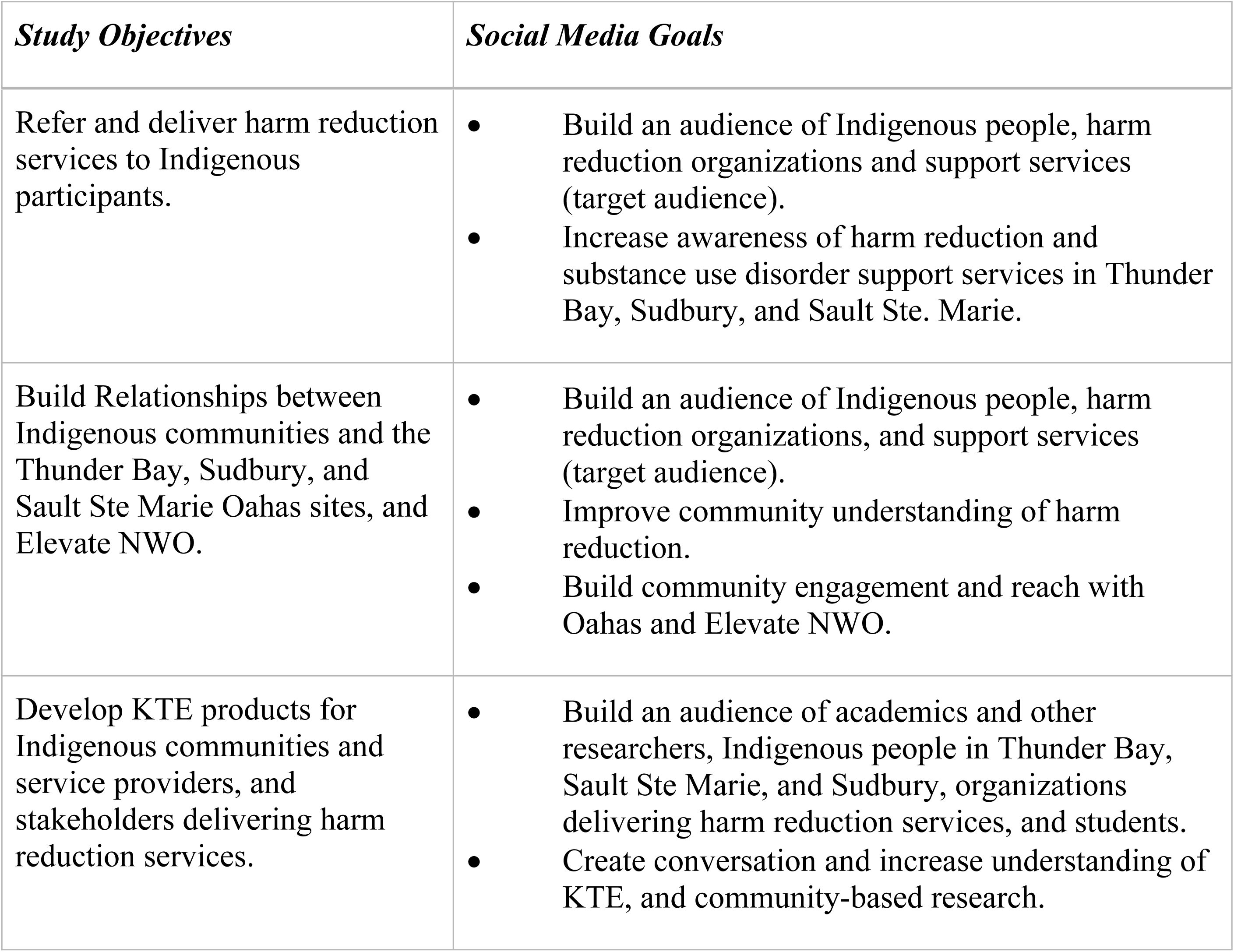
WHiSE 2.0 Objectives and Social Media Goals.

| *Study Objectives* | *Social Media Goals* |
| --- | --- |
| Refer and deliver harm reduction services to Indigenous participants. | • Build an audience of Indigenous people, harm reduction organizations and support services (target audience).<br>• Increase awareness of harm reduction and substance use disorder support services in Thunder Bay, Sudbury, and Sault Ste. Marie. |
| Build Relationships between Indigenous communities and the Thunder Bay, Sudbury, and Sault Ste Marie Oahas sites, and Elevate NWO. | • Build an audience of Indigenous people, harm reduction organizations, and support services (target audience).<br>• Improve community understanding of harm reduction.<br>• Build community engagement and reach with Oahas and Elevate NWO. |
| Develop KTE products for Indigenous communities and service providers, and stakeholders delivering harm reduction services. | • Build an audience of academics and other researchers, Indigenous people in Thunder Bay, Sault Ste Marie, and Sudbury, organizations delivering harm reduction services, and students.<br>• Create conversation and increase understanding of KTE, and community-based research. |

In all, the WHiSE 2.0 Social Media Strategy took place between the months of May 2024 and August 2024. While the planning phase occurred between May 1^st^ and May 24^th^, the doing phase took place between May 24^th^ and August 18^th^, and lastly, the evaluation phase took place in the last week of May, June, and July. Evaluation also occurred in the week of August 18^th^.

During the planning phase, a work plan was developed to ensure the successful dissemination of knowledge through social media. The plan highlighted the necessary activities needed to establish a successful knowledge translation strategy. In addition, it described the human and material resources imperative for the development of the Social Media Knowledge Translation Strategy, databases needed for literature reviews, helpful graphic design tools, and other software packages for knowledge dissemination. Moreover, it outlined deadlines and timelines for the completion of KTE activities.

As seen in Table 2, a social media engagement plan was also developed to ensure the successful dissemination of knowledge through social media. The goal of this plan was to emphasize the objectives of the WHiSE 2.0 project, establish specific objectives for social media, define measurable metrics to track the progress of these objectives, and devise a comprehensive strategy to attain them. In addition, the plan selected the social media platforms for the Social Media Knowledge Translation Strategy. As Table 3 illustrates, these platforms included Facebook, Instagram, and Twitter. For each, the engagement plan assessed their strengths and weaknesses, described the target audience, elucidated the types of posts to be shared, and specified the frequency of posts for optimal engagement. As Lu et al. indicated, one facet of good practices for harnessing social media for scholarly discourse, knowledge translation, and education, is understanding the nuances of specific platforms within the social media space [23].

**TABLE 2.**
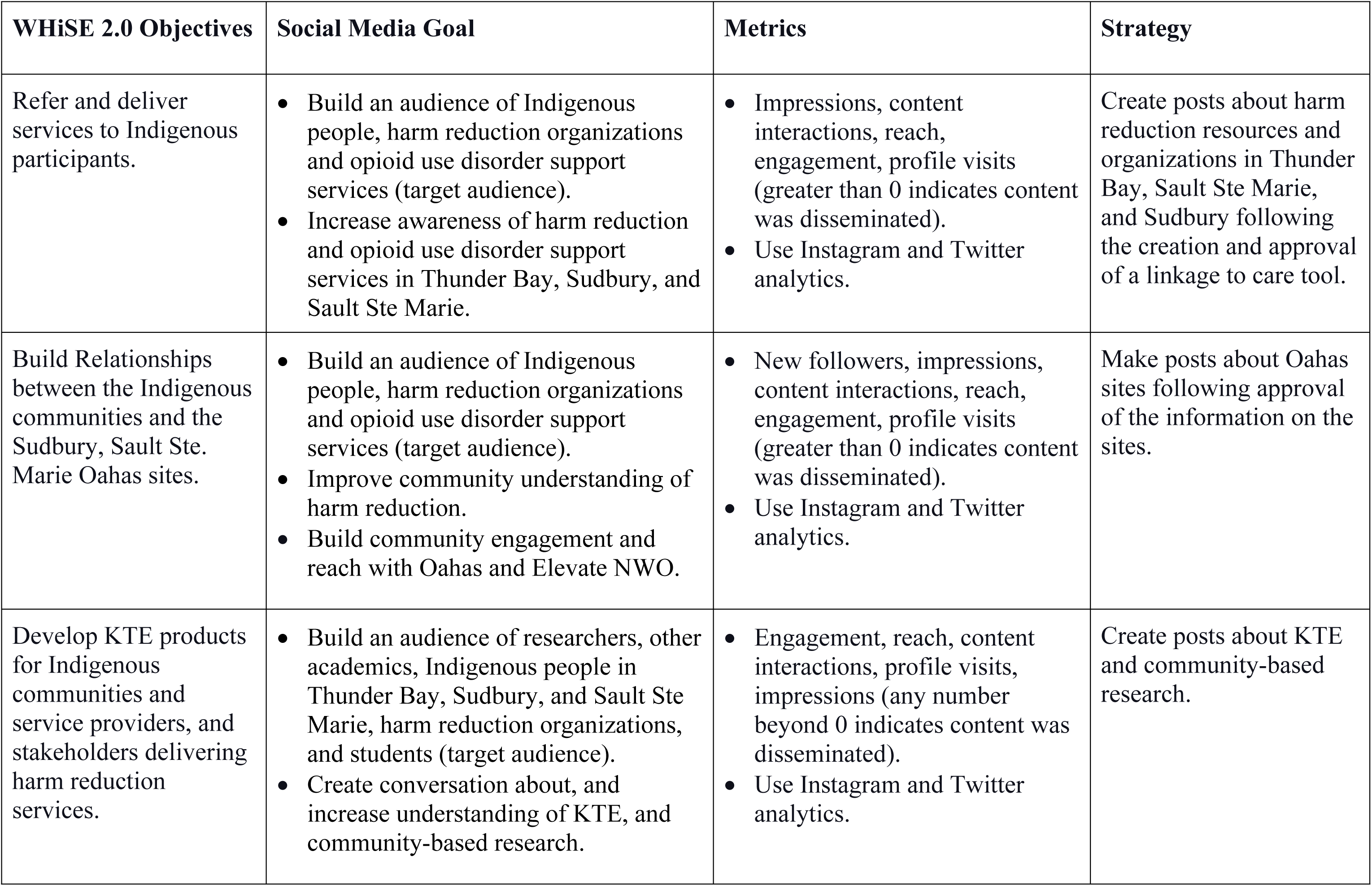
Social Media Engagement Plan Aligning WHiSE 2.0 Objectives with Social Medial Goals.

**TABLE 3.** Social Media Platforms.

| **Platform** | **Purpose** | **Type of Content** | **Target Audience** | **Specific Post** | **Frequency of Posts** |
| --- | --- | --- | --- | --- | --- |
| Facebook | • Spread information and educate target audience.<br>• Enables people to respond, share, and be updated on information quickly. | Informative and engaging posters, slideshows, infographics, and short videos | • Indigenous people in Thunder Bay, Sault Ste Marie, and Sudbury who access harm reduction services.<br>• Community-based organizations and service providers interested in harm reduction among Indigenous people.<br>• Students, researchers, and other academics and stakeholders. | • KTE posts<br>• Harm reduction resources | • Harm reduction: once a week<br>• KTE: biweekly |
| Instagram | • Stories: short video content to educate and capture attention of the audience (e.g., can be used to draw attention to main Instagram posts).<br>• Posts: Inform, educate, and engage. | Informative and engaging posters, slideshows, infographics, and short videos | • Indigenous people in Thunder Bay, Sault Ste. Marie, and Sudbury who access harm reduction services.<br>• Community-based organizations and service providers interested in harm reduction among Indigenous people.<br>• Students, researchers, and other academics and stakeholders. | • KTE posts<br>• Harm reduction resources | • Harm reduction: once a week<br>• KTE: biweekly |
| Twitter | • Conversations on specific topics.<br>• Connecting with others interested in study topics (e.g., using hashtags).<br>• Ensures that people are quickly informed and up-to-date. | Informative and engaging posters, slideshows, infographics, and short videos | • Indigenous people in Thunder Bay, Sault Ste Marie, and Sudbury who access harm reduction services.<br>• Community-based organizations and service providers interested in harm reduction among Indigenous people.<br>• Students, researchers, and other academics and stakeholders. | • KTE posts<br>• Harm reduction resources | • Harm reduction: once a week<br>• KTE: biweekly |

During the doing phase, infographics, videos, and posters were created and posted on Facebook, Instagram, and Twitter. This was accomplished with the use of CANVA, Sketch, and PowerPoint. In addition, individuals and organizations who were recognized as part of the target audience were followed on the three social media platforms. The doing phase began after the completion of the planning phase.

Lastly, during the evaluating phase, monthly twitter and Instagram analytics were used to assess engagement, reach, profile visits, new followers, and impressions. This phase occurred concurrently with the doing phase.

Content created for Facebook, Instagram, and Twitter included videos, posters, slideshows, and infographics. Each platform targeted Indigenous people in Sudbury, Sault Ste Marie, and Thunder Bay as well as community-based organizations delivering harm reduction, students, researchers, and academics. While information on harm reduction was disseminated weekly, information on KTE was shared on a biweekly schedule.

### Engaging in Indigenous KTE

Indigenous knowledge products must be disseminated in culturally appropriate ways. Thus, in creating Indigenous knowledge products, peer-reviewed research was obtained from Indigenous scholars and their allies. Further, culturally respectful language and imagery were used in Indigenous-centered content. Such imagery is illustrated in Figure 1.

**FIGURE 1.**
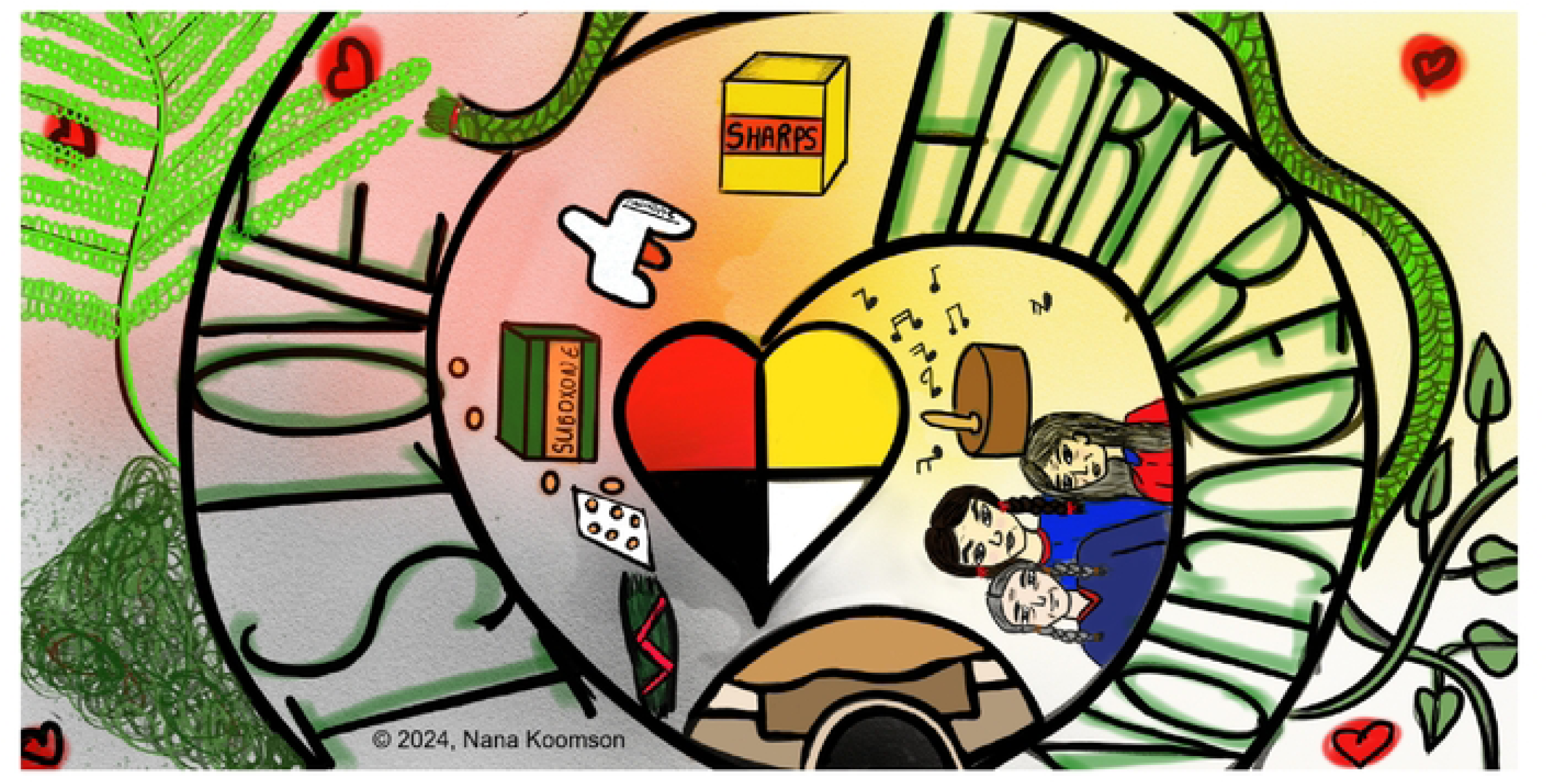
‘Harm Reduction is Love’ Rough Sketch Using Indigenous and Western-biomedicine Centered Images.

**FIGURE 2.**
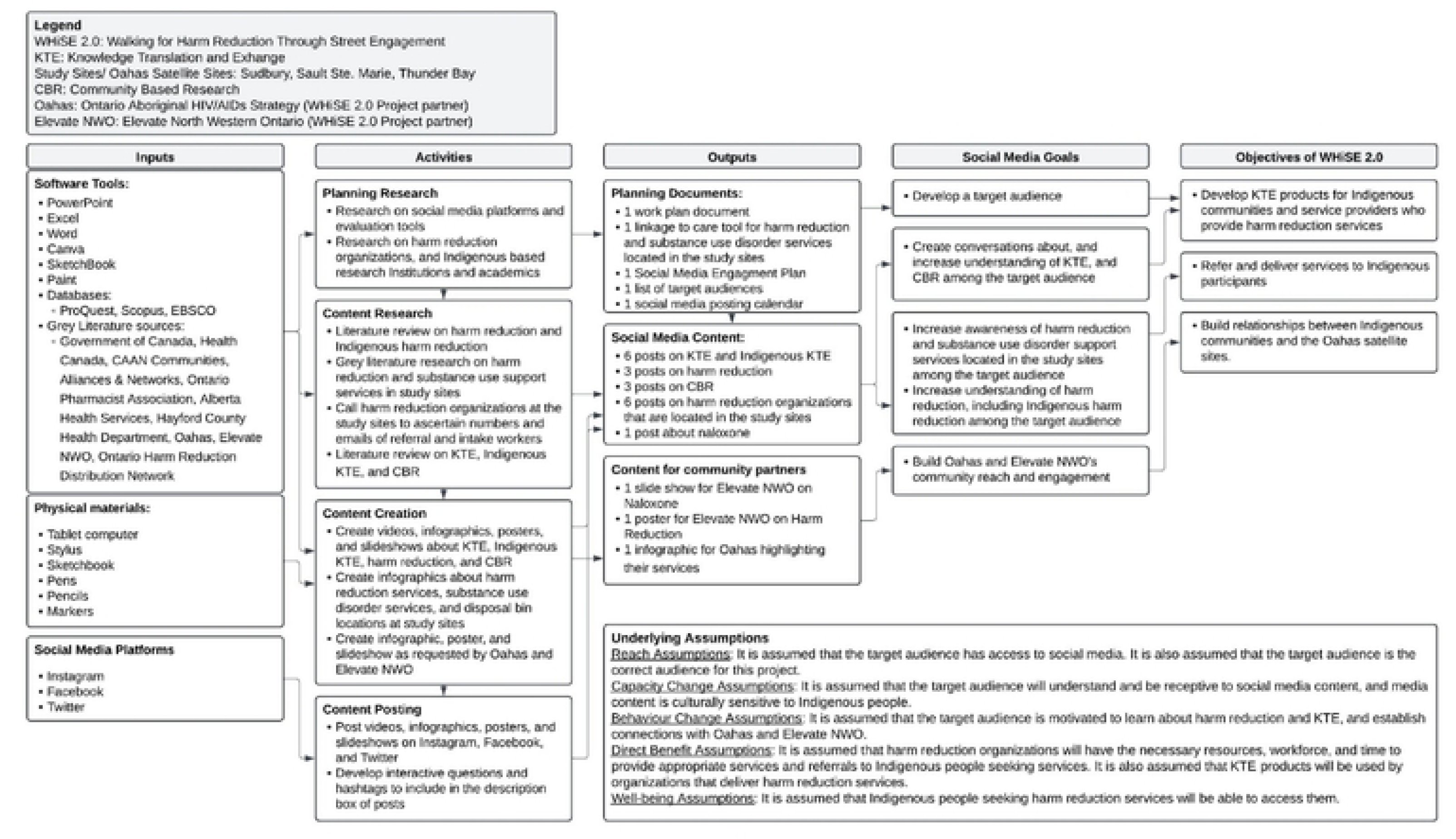
Logic Model of Social Media Knowledge Translation Strategy.

### Interacting With Community

To ensure that Indigenous KTE materials were culturally relevant and respectful, a significant aspect of the Social Media Knowledge Translation Strategy involved engaging with Indigenous communities. Notably, weekly meetings were held with an Indigenous Research Assistant. These sessions facilitated discussions on Indigenous teachings and ways of knowing, particularly in relation to harm reduction, and contributed to the development of media materials, as evidenced in Appendix I. Additionally, monthly discussions were held with both Elevate NWO and Oahas to discuss the progress of the WHiSE 2.0 study. These meetings were also an opportunity to address KTE requirements and explore ways to align KTE efforts with their specific needs. Lastly, a meeting was convened with Elevate NWO, Oahas, and an Indigenous Elder to discuss progress on the WHiSE 2.0 project, including the Social Media Knowledge Translation Strategy. During this session, the media products created for the strategy were presented.

### Develop a Target Audience (Reach 150 followers across all platforms)

#### Planning

The Social Media Knowledge Translation Strategy specifically targeted Indigenous individuals residing in Thunder Bay, Sault Ste. Marie, and Sudbury who engage with harm reduction services. Additionally, the strategy aimed to reach community-based organizations and service providers interested in Indigenous-focused harm reduction, students, researchers, and academics. To ensure outreach to this audience, a comprehensive list was compiled (n=26) using the Google search engine that included relevant educational institutions such as Algoma University, research institutes specializing in Indigenous-based research such as the University of Toronto’s Waakebiness Institute for Indigenous Health, Indigenous organizations such as the N’Swakamok Native Friendship Centre, Harm Reduction Organizations such as Elevate NWO, and academics involved in harm reduction research or research for racialized people. Team members like the Executive Director of Elevate NWO and Oahas members, specifically the Provincial Lead for Research & Evidence Integration, also provided valuable insights on organizations to follow.

#### Doing

To build an audience for the Social Media Knowledge Translation Strategy, targeted individuals, and organizations that adhered to the inclusion criteria described in the planning stage were followed on Twitter, Facebook, and Instagram. Furthermore, research was conducted on harm reduction, community-based research, KTE, and Indigenous KTE to create informative social media posts. This ensured the delivery of accurate and up-to-date information to the target audience.

### Increase Awareness of Harm Reduction and Opioid Use Disorder Support Services in Thunder Bay, Sudbury, and Sault Ste. Marie

#### Planning

A Linkage to Care tool was created to indicate the harm reduction and substance use disorder support services in Thunder Bay, Sault Ste. Marie, and Sudbury (Appendix II). This tool kit detailed the range of services provided at these sites, their emails, phone numbers, websites, physical locations, and hours of operation. The Linkage to Care tool also highlighted disposal bin locations in Thunder Bay, Sault St. Marie, and Sudbury, along with guidelines on acceptable items for disposal (Appendix III). Information used in the toolkit was derived from the Ontario Harm Reduction Distribution Program website [24]. Direct communication was also established with organizations to gather the contact details for referral and intake workers.

#### Doing

Using Canva, 6 infographics were created from the Linkage to Care tool. Each infographic featured 1 of the 3 study sites and included all the information outlined on the Linkage to Care tool. The infographics were shared on Instagram, Facebook, and Twitter in accordance with the social media posting calendar. Additionally, Elliot et al. described hashtags as a filtering technique that categorizes messages and leads individuals to conversations and discussions that pertain to specific topics or themes. Thus, hashtags such as #thunderbay, #saultstemarie, #sudbury, #harmreduction, and #opioidcrisis were employed.

### Improve Community Understanding of Harm Reduction

#### Planning

In Lu et al.’s 2021 paper, the authors highlight the lack of rigor during the production and use of social media resources as a pitfall to social media knowledge translation. Thus, a search strategy was developed to aid in a literature review of harm reduction principles, and Indigenous harm reduction strategies. Both peer-reviewed and grey literature sources were consulted. The search process included terms such as ‘Harm Reduction,’ ‘Indigenous,’ ‘opioid agonist treatment,’ ‘safe consumption site,’ ‘Good Samaritan Drug Overdose Act,’ and ‘needle exchange.’ Peer-reviewed studies were obtained from reputable databases such as ProQuest, Scopus, and EBSCO, while insights from grey literature were sourced from CAAN Communities, Alliances, & Networks (formerly known as the Canadian Aboriginal AIDS Network) [25] and Health Canada [26].

#### Doing

Following research, CANVA, Paint, Sketch, and PowerPoint were used to create 2 visually appealing slideshows and 1 video that focused on harm reduction topics. Images were either drawn or obtained from CANVA or PowerPoint. Moreover, with consideration to lay audiences, language and sentence structures were tailored to a reading level appropriate for grades 5 to 8 [23]. The slideshows and video were posted on Instagram, Facebook, and Twitter according to the social media posting calendar. In addition, a description was curated for each post that included questions such as ‘Have you heard of the Good Samaritan Drug Overdose Act?’ ‘What Harm Reduction Services do you know?’ and ‘Have you heard of the Principles of Harm Reduction?’ Finally, hashtags such as #harmreductionsaveslives, #harmreduction, #opioidcrisis, #goodsamaritandrugoverdoseact, #indigenoushealth, and #indigenousharmreduction were used.

### Build Community Engagement and Reach with Oahas and Elevate NWO

#### Planning

Using the Oahas website, a search was completed to gather information on their services [27]. Additionally, following conversations with Elevate NWO about the importance of disseminating information about Naloxone safety, specific sites were reviewed for information pertaining to naloxone safety, uses, and storage. Grey literature sources for this search include the Government of Canada [28], the Ontario Pharmacist Association [29], Alberta Health Services [30], and Hayford County Health Department [31].

#### Doing

Using Canva and information from the grey literature review, an infographic was created and shared with stakeholders within Oahas. Images were obtained from Canva and the Oahas website. Again, upon the request of stakeholders within Elevate NWO, a slideshow was created on naloxone and shared with Elevate NWO stakeholders. The slideshow was also posted on Instagram, Facebook, and Twitter. Hashtags that were used included #naloxone, #naloxonesaveslives, #harmreduction, and #opioidcrisis. Finally, following consultation with Elevate NWO stakeholders, a sketch was created using SketchBook. As illustrated in Figure 1, the Sketch highlighted the topic of ‘Harm Reduction is Love’ and featured imagery that pertained to Indigenous-specific harm reduction, and Western biomedical perspectives of harm reduction, The sketch was shared with Elevate NWO and Oahas stakeholders.

### Create Conversation and Increase Understanding of Knowledge Translation and Exchange, and Community-Based Research

#### Planning

A search strategy was developed to aid in a literature review of KTE, Indigenous KTE, foundations of community-based research, and community-based research principles. Peer-reviewed literature sources were consulted. The search process included terms such as ‘Knowledge translation and exchange,’ ‘Indigenous knowledge translation,’ ‘knowledge dissemination,’ ‘knowledge sharing,’ ‘community-based research,’ and ‘community-based participatory research.’ Peer-reviewed articles were obtained from databases such as ProQuest, Scopus, and EBSCO. The search took place between May 5^th^, 2023, and May 9^th^, 2023 and only included articles written in English. Lastly, this was not a comprehensive search, and databases were selected from popular databases accessible through the University of Toronto’s e-library.

#### Doing

Using information from the literature review, 8 videos, and 1 poster was created using Canva, PowerPoint, and Paint. Images were either drawn or obtained from CANVA or PowerPoint. Moreover, considering lay audiences, language and sentence structures were tailored to a grade 5 to 8 reading level [23]. The videos and poster were posted on Instagram, Facebook, and Twitter according to the social media posting calendar. In addition, a description was created for each post that included questions such as ‘How has knowledge been shared with you?’ Have you pushed or pulled research before?’ and ‘Have you heard of the Principles of Harm Reduction?’ Finally, hashtags such as #kte, #knowledgetransfer, #push, #pull, #integratedknowledgetranslaton, #collaboration, #communitybasedresearch, and #participatoryresearch were used.

### Evaluating

Monthly Twitter analytics and Instagram insights were used to access metrics on engagement rate and profile visits. Instagram metrics that were collected included: the number of new followers, engagement, reach, and profile visits. Metrics collected from Twitter analytics included: the number of new followers, impressions, and profile visits. These metrics were used to analyze the success of the Social Media Knowledge Translation Strategy. While monthly metrics were collected, in the last month of observations, data was only collected for 17 days. Facebook insights were not collected or monitored. However, as the Instagram page was linked to a Facebook page, content posted on Instagram was automatically posted on Facebook.

### Underlying Assumptions

The development of a Social Media Knowledge Translation Strategy for the WHiSE 2.0 project is grounded in several underlying assumptions related to reach, capacity change, behavior change, direct benefit, and well-being. For the strategy’s objectives to be realized, these assumptions must hold true [32].

Regarding ‘reach’, it is expected that the target audience has access to social media and will actively engage with the social media accounts designated for this Social Media Knowledge Translation Strategy. Further, it is assumed that the identified target audience is the appropriate audience for KTE products, service referrals, and the promotion of Oahas and Elevate NWO.

In terms of ‘capacity change’, it is presumed that the target audience will comprehend, be receptive to, and engage with the content shared on the selected social media platforms. It is also assumed that the content posted on WHiSE social media accounts is culturally sensitive to Indigenous communities and fosters a respectful and inclusive approach to communication.

For ‘behavior change’ assumptions, it is expected that the target audience is motivated to acquire knowledge about harm reduction services and organizations, establish connections with Oahas and Elevate NWO, and learn about KTE. Furthermore, it is presumed that creating social media posts about KTE and harm reduction will contribute to the improved delivery and referral of harm reduction services, stronger relationships with Oahas and Elevate NWO, and increased awareness of KTE.

In terms of ‘direct benefit’, it is assumed that harm reduction organizations will have the necessary resources, workforce, and time to provide appropriate services and referrals to Indigenous individuals seeking assistance. Additionally, it is anticipated that the KTE products will be used by community-based organizations interested in delivering harm reduction services to Indigenous communities.

Lastly, in consideration of ‘well-being’ assumptions, it is assumed that Indigenous individuals seeking harm reduction services can access harm reduction organizations. Such access can include affordable transportation options. For instance, in Davy et al.’s 2016 paper on access to primary health care services for Indigenous peoples, the authors assert that the cost of transport to health care services was as prohibitive as the cost of health care for Indigenous people. As such, some Indigenous health care service providers offer transportation to and from their facility or provide outreach services that deliver care services to a client’s home [33].

### Theory of Change

If a Social Media Knowledge Translation Strategy is implemented to disseminate knowledge about KTE and harm reduction to the specified target audience, and if the underlying assumptions hold, then the 3 KTE-specific objectives of the WHiSE 2.0 study will be met. Thus, there will be effective referral and delivery of harm reduction services, established relationships between Indigenous communities and harm reduction organizations, and increased knowledge of KTE and KTE strategies among the target audience [32].

### Development of a Logic Model

A logic model was crafted using CANVA. Throughout this initiative, inputs and activities were meticulously recorded through the structured workplan. The outcomes derived from these inputs and activities were subsequently correlated with the social media goals, which, in turn, were aligned with the overarching objectives of the WHiSE 2.0 study. In developing the logic model, it was assumed that a coherent sequence exists between the inputs and activities within the social media strategy and the goals of the WHiSE 2.0 study [34].

## RESULTS

Five planning documents were created for this Social Media Knowledge Translation Strategy, including a work plan, a linkage to care tool for harm reduction and substance use disorder services, a social media engagement plan, a list of target audiences (n=26), and a social media posting calendar. The 5 planning documents were consulted in the creation of 21 media products. This included 9 videos, 2 posters, 7 infographics, and 3 slideshows. 19 of these media products (9 videos, 1 poster, 6 infographics, 3 slideshows) were posted on Instagram, Facebook, and Twitter. Based on WHiSE 2.0 objectives, and the social media goals, a logic model resulted as a visual representation of this social media knowledge translation approach. The logic model highlights the resources needed to complete the Social Media Knowledge Translation Strategy for WHiSE 2.0, the activities undertaken during the planning and doing stage of the Social Media Knowledge Translation Strategy, and the planning documents and media products that resulted.

### Target Audience

An essential aspect of the Social Media Knowledge Translation Strategy revolved around effectively reaching the target audience. In addition to a dedicated list (n=26) of academics, research institutions, and harm reduction organizations, relevant social media handles outside the curated target audience list were followed. Actively following and interacting with accounts outside the targeted list led to an increased following, reach, and engagement on all social media platforms.

Most of the followers on Twitter and Instagram were academics, publication journals, Indigenous organizations, harm reduction organizations, and students. Thus, the desired target audience was reached. While this was the case, the platforms did not reach 150 followers by the end of the data collection period, which was from May 2024 to August 2024. While Instagram reached 55 followers, Twitter reached 37 followers, and Facebook reached 7 followers.

### Instagram and Twitter Analytics

As is seen in Table 4, results from Instagram Insights revealed that over the course of this project, there was an average of 95 content interactions and 126 profile visits per month. However, the average number of new followers was 7 per month. Moreover, each month, there was an average of 16 engagements and 32 reaches from followers, and 11 engagements and 136 reaches from non-followers. The Instagram account was established in February 2023, and data was collected from May to August 2023.

**TABLE 4.**
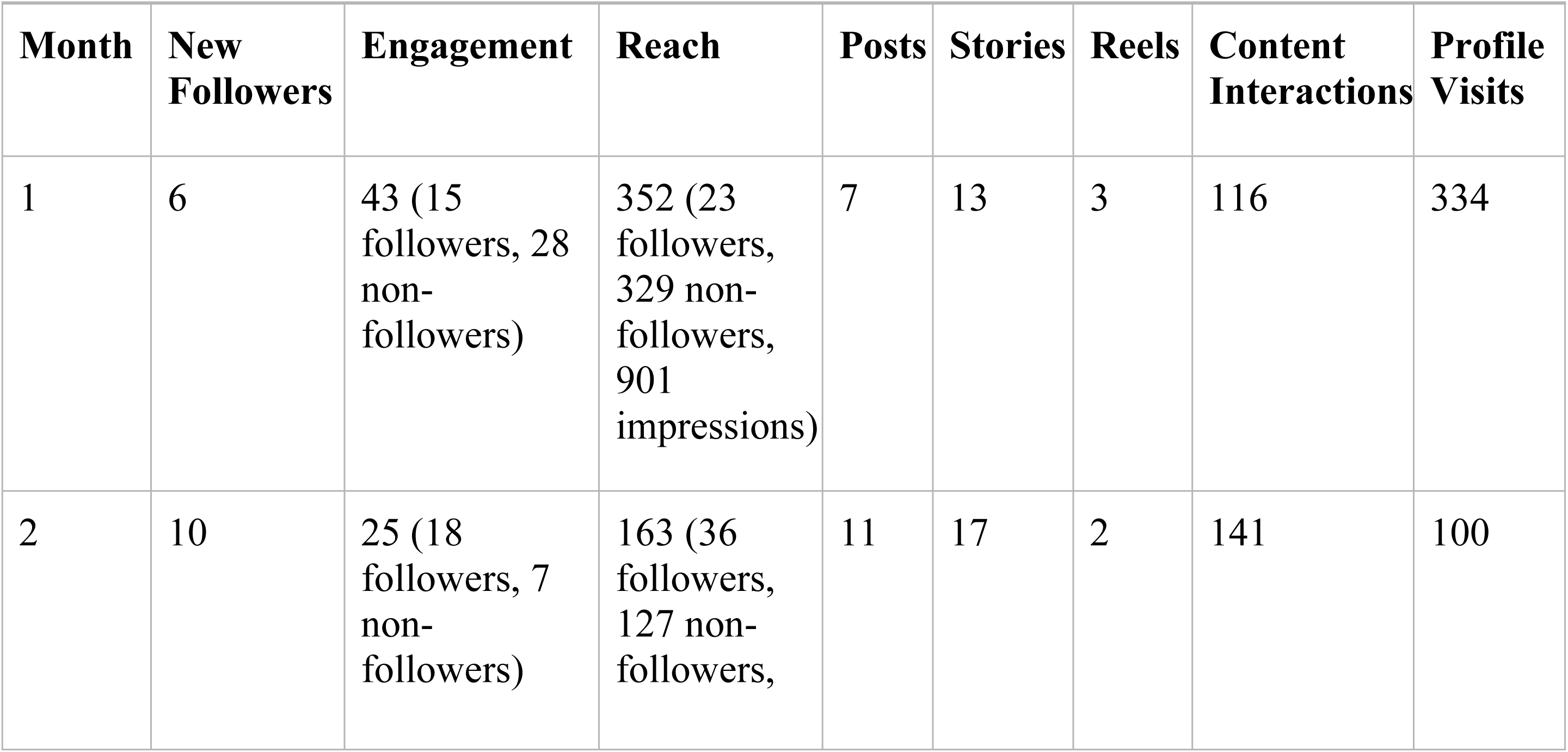
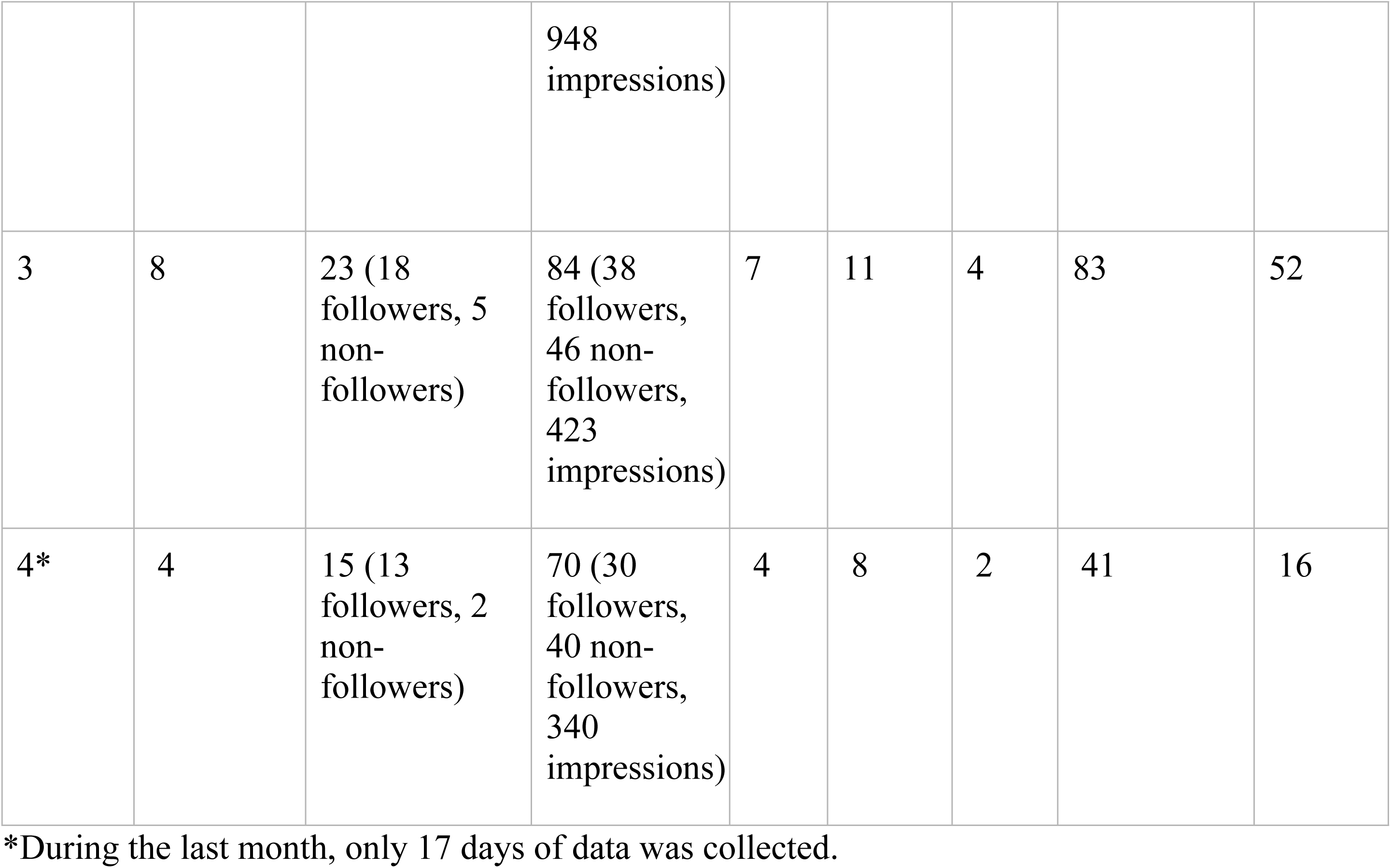
Instagram Metrics.

| Month | New Followers | Engagement | Reach | Posts | Stories | Reels | Content Interactions | Profile Visits |
| --- | --- | --- | --- | --- | --- | --- | --- | --- |
| 1 | 6 | 43 (15 followers, 28 non-followers) | 352 (23 followers, 329 non-followers, 901 impressions) | 7 | 13 | 3 | 116 | 334 |
| 2 | 10 | 25 (18 followers, 7 non-followers) | 163 (36 followers, 127 non-followers, | 11 | 17 | 2 | 141 | 100 |
|  |  |  | 948 impressions) |  |  |  |  |  |
| 3 | 8 | 23 (18 followers, 5 non-followers) | 84 (38 followers, 46 non-followers, 423 impressions) | 7 | 11 | 4 | 83 | 52 |
| 4* | 4 | 15 (13 followers, 2 non-followers) | 70 (30 followers, 40 non-followers, 340 impressions) | 4 | 8 | 2 | 41 | 16 |

\*During the last month, only 17 days of data was collected.

Twitter Analytics (Table 5) indicate that there was an average number of 4 new followers per month and an average number of 1,661 impressions per month. Again, in months 2 and 3, there was an average of 451 profile visits per month. The Twitter account was established in February 2023, and data was collected from May to August 2023.

**TABLE 5.**
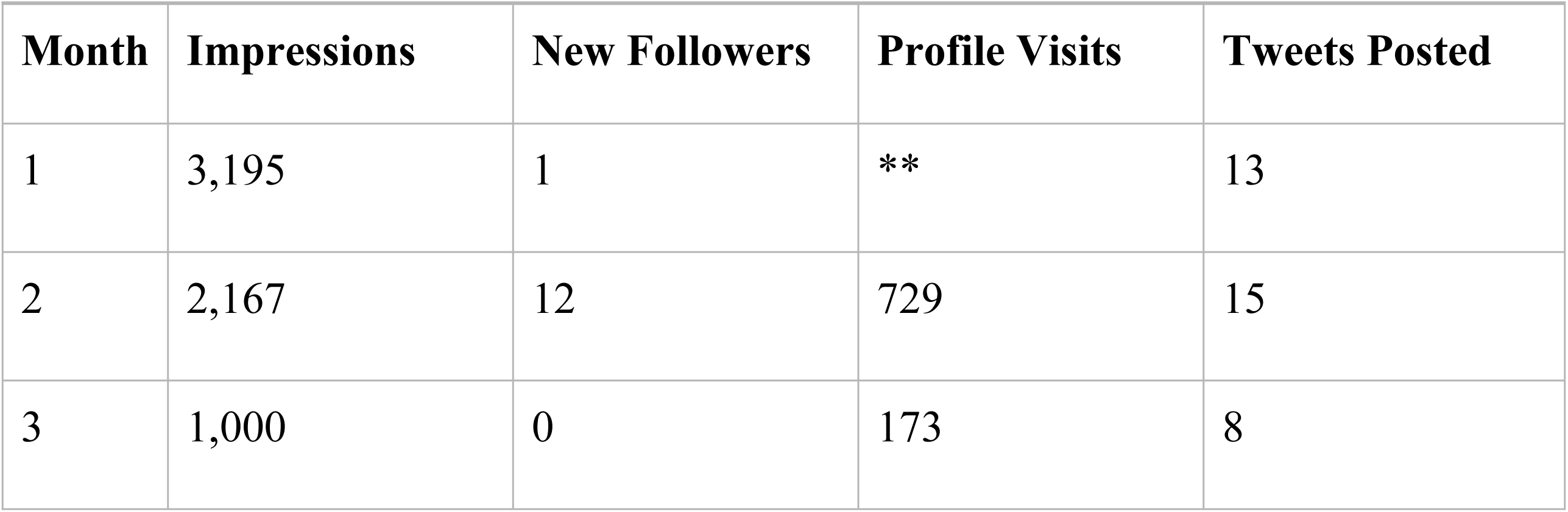
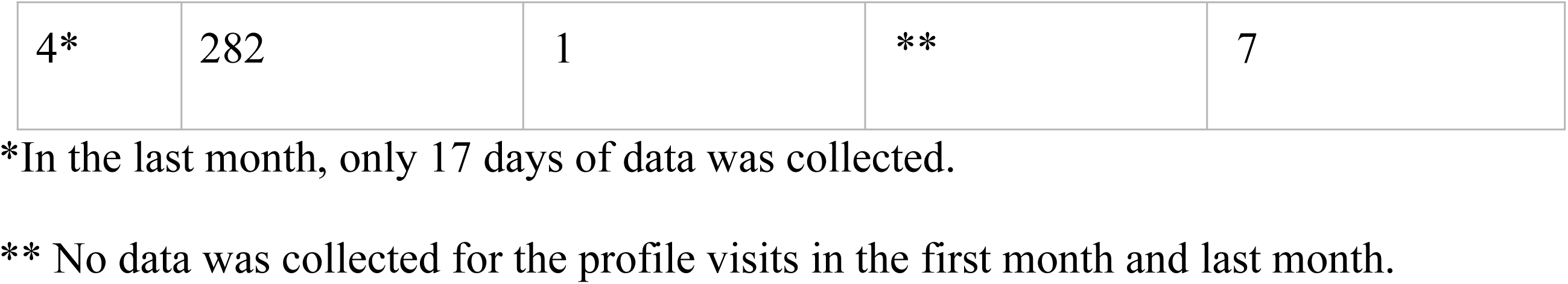
Twitter Metrics.

| Month | Impressions | New Followers | Profile Visits | Tweets Posted |
| --- | --- | --- | --- | --- |
| 1 | 3,195 | 1 | ** | 13 |
| 2 | 2,167 | 12 | 729 | 15 |
| 3 | 1,000 | 0 | 173 | 8 |
| 4* | 282 | 1 | ** | 7 |

\*In the last month, only 17 days of data was collected.
\*\* No data was collected for the profile visits in the first month and last month.

As is seen from Tables 4 and 5, there was a disproportionate ratio of new followers to reach, content interactions, and profile visits. Additionally, there was higher reach and impressions compared to engagement. Thus, there were more unique users who saw the knowledge products (reach) and more impressions (the number of times content was viewed) than followers. These observations indicate that while the content reached a wide audience, it remains uncertain whether this audience was part of the intended target audience. The discrepancy in the number of followers to non-followers concerning reach further emphasizes this concern (on average, there was a 1:4.3 ratio of followers to non-followers per month). It is also visible in the number of new followers to profile visits and content interactions which were 1:18 per month, and 1:13.6 per month respectively on Instagram.

## DISCUSSION

### Summary of Results

This project aimed to create a Social Media Knowledge Translation Strategy for WHiSE 2.0 that targeted Indigenous audiences and their stakeholders in Thunder Bay, Sault Ste. Marie, and Sudbury. The strategy highlighted elements of Indigenous knowledge translation, harm reduction, and community-based research using the reach of mass media.

As seen in Table 2, any quantity of impressions, profile visits, content interactions, reach, engagement, and new followers beyond 0, was analyzed as successfully disseminating knowledge to at least 1 organization or individual. Based on these standards, while 150 followers across all platforms was not realized, the goal of increasing awareness and conversation around the study topics was successfully accomplished. However, it is evident that additional efforts are needed to reach the intended target audience and encourage social media users who interact with the content or visit the project’s social media pages to become followers.

### Comparison to Other Studies

Previous research has highlighted the significance of social media as a valuable tool for health-seeking behavior, particularly among Indigenous populations. In a 2021 article by Carlson et al., Aboriginal and Torres Strait Islanders in Australia were found to use platforms like Twitter and Facebook as service directories to discover relevant and easily accessible health services [35]. These findings underscore the importance of social media posts promoting harm reduction services and organizations in Thunder Bay, Sault Ste. Marie, and Sudbury, despite a disparity between audience reach and engagement.

Similarly, social media has proven to be an effective medium for educating specific target audiences on critical public health matters. A quasi-experimental feasibility study by Gouh et al. evaluated the use of Twitter in a skin cancer prevention campaign. The campaign generated 417,678 tweet impressions, 11,213 engagements, and 1,211 retweets, increasing knowledge about skin cancer severity and raising awareness that, while skin cancer is the most common form of cancer, melanoma is the most serious [36]. As with this study, social media, particularly Twitter, was able to disseminate important health-related information that positively impacted public knowledge.

A 2019 study by Dehlin et al. also highlighted the potential of social media as a tool for raising awareness about public health interventions. Specifically, the researchers assessed whether the Prep4Love campaign succeeded in increasing awareness of Pre-exposure Prophylaxis (PrEP) among Black men who have sex with men and Black transgender women in Chicago. Similar to this research, the authors used platforms such as Instagram, Facebook, and targeted smart ads to promote the campaign. Their efforts successfully achieved the primary goal of disseminating PrEP information, generating a total of 40,913,560 unique views across the chosen social media channels [37].

Building on these examples, the effectiveness of social media in spreading harm reduction awareness is especially relevant to the public health messages in this study’s Social Media Knowledge Translation Strategy. In 2022, Ackermann et al. explored awareness, understanding, and perceptions of the Good Samaritan Drug Overdose Act among individuals likely to witness an overdose in British Columbia. Participants identified social media as a direct and effective channel for public health education, particularly regarding the Good Samaritan Drug Overdose Act, reaching a broad audience [38].

While Twitter and Instagram metrics within this project indicate more reach and impressions compared to engagement, and more non-followers reached compared to followers, other studies have highlighted that this is neither unusual nor negative [39–40]. For instance, a study focusing on Covid-19 messaging on social media for American Indian and Alaska Native communities found that campaign posts generated a significant 425,834 impressions but only garnered 6,016 engagements, resulting in an average engagement rate of 2.2% [41]. Interestingly, as the one-year campaign progressed, the average number of impressions per post increased, while the average engagement rate declined. This trend indicated that despite a growing follower count, individuals were becoming less inclined to actively share or amplify the campaign’s message [41]. Other studies have similarly identified trade-offs between impressions, reach, and engagement [40–41]. In a study using Facebook to promote HPV vaccination coverage among Danish girls, researchers discovered that although the total reach of posts was 3,476,023 people, the highest engagement rate among 84 different posts was only 15% [40]. The authors cautioned that engagement rates alone do not convey the sentiment, positive or negative, associated with engagement. Consequently, achieving a high engagement rate may not necessarily be deemed successful if it comes at the expense of a lower overall reach [40]. This further highlights the success of this social media strategy, despite the low engagement rates.

In fact, a study conducted in 2019 delved into the health-related content shared on social media among Aboriginal and Torres Strait people in Australia. The authors categorized social media users into different roles, including observers, post-sharers, positive supporters, educators, and influencers [39]. Analysis based on these roles revealed that a substantial portion of the audience for this project primarily assumed the role of observers. According to Hefler et al., observers typically refrain from liking or sharing content but occasionally consume content that captures their interest [39]. This study also involved a smaller proportion of positive supporters who actively engaged with posts. Hefler et al. note that the engagement of positive supporters is more subtle and often involves liking a post to express approval [39].

### Indigenous Knowledge Translation and Exchange

There are limited descriptions of Indigenous KTE. Some scholars have described it as an Indigenous-led sharing of culturally relevant health information and practices to improve Indigenous health, policy, services, and programs [42–43]. All aspects of the research process including knowledge dissemination with and for Indigenous people must uphold the principles of reciprocity, respect, relevance, responsibility, reverence as well as the OCAP™ principles which includes ownership of data, control over how information is used and accessed, and stewardship of information [3, 42, 44].

Further, it has been illustrated by some authors, that dissemination products must fall within the priorities and ways of knowing identified by Indigenous carriers and translators of knowledge, and knowledge users [42, 45]. These dissemination products are shared in diverse ways that include orality, drawings, songs, paintings, dance, and theatre [46]. Consequently, generalized knowledge translation activities are conceptualized by some Indigenous scholars as reflective of colonized practices because these activities often assume what is best for Indigenous people [47].

Wright et al. have described Indigenous KTE as being composed of 5 methodological features. These include drawing on Indigenous ways of knowing, employing a decolonized approach, supporting self-determination, grounding the process in participatory research approaches, and creating an ethical space for research dissemination where dialogue between Indigenous and Western knowledge systems, cultures, and worldviews can collaborate without hierarchy [47].

Social media can be an effective tool for applying the principles and methodological features of Indigenous KTE. Since knowledge translation activities that link health research to practice in Indigenous communities are often overlooked, it has been suggested that social media knowledge translation can help bridge this gap [21–22]. In this study, the Social Media Knowledge Translation Strategy aimed to reflect Indigenous KTE principles by disseminating culturally relevant and respectful imagery on Facebook, Instagram, and Twitter, with input from an Indigenous Research Assistant. However, the limited research on Indigenous KTE posed a challenge, as there were few examples to guide the development of our strategy.

### Limitations and Future Directions

In addition to limited research on Indigenous KTE, there are several other limitations to consider when interpreting the results of this study. Firstly, the evaluation period of 4 months may not have been sufficient to fully assess the effectiveness of the Social Media Knowledge Translation Strategy. A longer timeframe is required to build and reach a more substantial audience and establish meaningful relationships between the target audience and harm reduction organizations. This is especially evident in the limited time that was available to collect data in the last observation month. Also, search strategies used for literature reviews focused on databases. As such, some Indigenous-specific journal which are not indexed may not have been captured. Again, it is necessary to acknowledge the limitation of online metrics as an exclusive indicator of meeting study objectives. Online metrics, such as reach, impressions, engagement, and content interactions, serve as proxy indicators [48] and do not guarantee active attention from the audience toward the knowledge products. Elliot et al. also noted that measuring the success of a Social Media Knowledge Translation Strategy is complex and may require unconventional evaluation methods. It is particularly difficult to directly link knowledge sharing to behavioural changes or to assess its impact on health outcomes and health systems. While KTE strategies can enhance knowledge and awareness, and social media analytics can offer proxy indicators, their capacity to assess more long-term health outcomes remains limited [14]. Moreover, there are few standard metrics for social media engagement, and varying definitions and use [49]. Additionally, during the development and implementation of the strategy, community partners were interested in different aspects of the strategy. Consequently, some aspects of the strategy catered to their unique organizational needs which may have impacted the reach of the strategy. Lastly, most followers were initially followed by this project’s social media account based on the curated list of relevant accounts. As a result, not all possible community-based harm reduction and Indigenous organizations, academics, and researchers were followed. This may have limited the reach of the disseminated knowledge products.

Considering these limitations, future iterations of the Social Media Knowledge Translation Strategy should incorporate a longer evaluation period to assess its impact more effectively. Notably, in Loft et al.’s 2020 study, a period of 7 months was allocated to posting content on Facebook, while in Weeks et al.’s 2022 study, Twitter posts were published and analyzed over one year. In addition, expanding the range of metrics beyond Instagram and Twitter analytics is crucial. One such metric can involve assessing the increase in visits to harm reduction organizations in Thunder Bay, Sudbury, and Sault Ste Marie from individuals who first discovered these organizations from this project’s social media. Again, to address the disproportionate ratio of new followers to profile visits, future iterations of this strategy should consider making their social media pages more personable and relatable. Making relatable content outside education and service referrals may not only attract more new followers, but developing personalized content that resonates with the target audience can foster a stronger sense of connection and representation on the project’s social media pages. This, in turn, may lead to increased engagement with educational and referral content [50].

## CONCLUSION

The Social Media Knowledge Translation Strategy designed for the WHiSE 2.0 study aimed to bridge the knowledge-to-practice gap between Indigenous and harm reduction organizations, and harm reduction and KTE knowledge. While the targeted number of followers was not reached on all platforms, the desired target audience of harm reduction and Indigenous organizations, students, researchers, and other academics was curated. Moreover, knowledge products were successfully disseminated to at least one person using social media. In all, future iterations of the strategy should allocate more time to planning, doing, and evaluating. Moreover, other metrics beyond social media insights must be collected to fully evaluate the success of the strategy.

## Data Availability

All relevant data are within the manuscript and its Supporting Information files.

## ACKNOWLEDGMENTS

The authors would like to acknowledge the guidance of Elevate NWO and the Ontario Aboriginal HIV/AIDS Strategy for the development of the Knowledge Translation materials.

## APPENDIX 1

### Media Content

**Posters, Infographics, and Slideshow:**

https://www.anitacbenoit.com/copy-of-infographics

**Videos.**

https://www.anitacbenoit.com/videos

## APPENDIX II

*Linkage to Care Tool-Harm Reduction Resources and Services*

## APPENDIX III

*Linkage to Care Tool-Disposal Bin Locations*

